# The impact of COVID-19 pandemic on AMI and Stroke mortality in Lombardy: Evidence from the epicenter of the pandemic

**DOI:** 10.1101/2021.04.15.21255255

**Authors:** C. Rossi, P. Berta, S. Curello, P.G. Lovaglio, M. Magoni, M. Metra, A.M. Roccaro, S. Verzillo, G. Vittadini

## Abstract

**Background:** The Covid-19 pandemic has enormously impacted the delivery of clinical healthcare and hospital management practices in most of the hospitals around the world for both Covid and no-Covid patients. In this context, it is extremely important to assess whether the clinical management of no-Covid cases has not seriously been compromised during the first epidemic outbreak. Among no-Covid cases, patients with acute myocardial infarction (AMI) and stroke need non-deferrable emergency care and are the natural candidates as no-Covid patients to be studied. Preliminary evidence suggests that i) the time from onset of symptoms to emergency department (ED) presentation has increased in Covid-19 times as well 30-day mortality during the pandemic has been higher. We aimed to complement this evidence assessing if the additional stress due to the high inflow of Covid-19 patients at hospital level has modified AMI and Stroke admission criteria and related mortality rates in a causal inference framework.

**Methods:** To study the impact of Covid pandemic on mortality rates for AMI and Stroke we adopt two quasi-experimental approaches, regression-discontinuity design (RDD) and difference-in-regression-discontinuity (DRD) designs by which we identify the plausible causal effect on mortality of the Covid-19-related hospital stress due to the introduction of State of Emergency restrictions.

**Findings:** We check the causal effect of the Covid-19 pandemic on mortality rates of AMI and stroke over several time-windows of 15-days around the implementation date of the State of Emergency restrictions for COVID-19 (March, 9^th^). Despite the potential adverse effect on expected mortality due to a longer time to hospitalization, the AMI and Stroke mortality rates are overall not statistically different from the one observed in the control group. The obtained results provided by RDD and DRD models are robust also when we account for seasonality and unobserved factors.

**Interpretation:** In a quasi-experimental setting we assessed the causal impact of the hospital and staff extra-burden generated by the first wave of Covid-19 patients on mortality rates of no-Covid non-deferrable urgent cases (AMI and Stroke) hospitalized at Spedali Civili of Brescia, one of the most hit provinces in Italy by Covid-19 during March and May 2020. We find a non-statistically significant impact on mortality rates for AMI and Stroke patients providing evidence of the hospital ability to manage - with the implementation of a double track organization-the simultaneously delivery of high quality cares to both Covid and no-Covid patients. Availability of similar data for the regional context as a whole is needed to further substantiate the findings and explore existing differences in efficacy of different managerial settings implemented in Lombardy hospitals.

**Funding:** All authors - except for Stefano Verzillo-acknowledge financial support from the Region of Lombardy, project 2014IT16RFOP012 ‘Misura a sostegno dello sviluppo di collaborazioni per l’identificazione di terapie e sistemi di diagnostica, protezione e analisi per contrastare l’emergenza Coronavirus e altre emergenze virali del futuro’. Stefano Verzillo has participated as external econometrician from European Commission, Joint Research Center to this project without receiving any funding or financial support, in compliance with EC rules. His contribution has been offered in the Conceptualization and Writing and Editing stages.

**Role of the Funding source:** The founder had no roles in our study design, data collection and analysis, decision to publish and preparation of the manuscript.

## Introduction

The Covid-19 pandemic has enormously impacted the delivery of clinical healthcare and hospital management practices in most of the hospitals around the world. If on the one hand an extraordinary effort has been exerted to face the workload due to hospital admissions of Covid-19 patients on the other hand it is extremely important to assess whether the clinical management of no-Covid cases has not seriously been compromised during the first unexpected epidemic outbreak.

Around the world, no-Covid hospital admissions fell precipitously with the declaration of the coronavirus disease 2019 (Covid-19) pandemic (Birkmeyer et al 2020). Volumes declines for elective surgery or non-critical patients’ medical services, as well as for acute cases like strokes and acute myocardial infarctions (Arcaya et al 2020; De Rosa et al 2020; Mahmud et al 2020; Siegler et al 2020; Solomon et al. 2020), have been registered all over the world during the Covid-19 pandemic.

Declining hospitalization rates may indicate that patients defer care for life-threatening conditions with substantial damage to public health. Moreover, changes in admissions rates may differ by medical condition/diagnosis and ultimately by illness severity which may reflect in significant changes of in-hospital mortality rates.

It can be reasonably stated that if Covid-19 has modified hospitalization selection criteria (cutting hospitalization for less seriously ill patients), we expected similar pattern on (raising) in-hospital mortality rates, unless organizational changes implemented to face the pandemic have ensured appropriate levels of clinical care for patients with no-Covid clinical conditions that require hospital care.

It has been stated that no-Covid patients are not seeking hospital admissions because of their concerns about the risk of nosocomial COVID-19 infection, as well as because of the social limitations put in place from governments and local health authorities to face the pandemic (Abdelaziz et al 2020; Huet et al, 2020).

To this aim, monitoring the effective systematic changes at hospital level during the Covid-19 pandemic is crucial for all the National Health Systems (NHS).

To assess this issue, literature has focused on a particular subgroup of hospital admissions. To get rid off the selection bias affecting most of the no-Covid diseases patients with acute myocardial infarction (AMI) and stroke - needing non-deferrable emergency cares-are the natural candidates’ groups of no-Covid patients to be studied.

Recent literature shows how - for urgent diseases-the time from onset of symptoms to emergency department (ED) presentation has increased in Covid-19 times in Lombardy Region (Gramegna et al. 2020; Mitra et al 2020). In addition, recent evidence from UK show how the pool of admitted patients for AMI was easier to be admitted in hospital (due to a younger and less severe case-mix) and, for NSTEMI AMI, had higher 30-day mortality during the pandemic (Wu et al. 2020, Mitra, Biswadev, et al. 2020, Rudilosso et al. 2020).

Finally, a recent multicenter observational report from Italy found that AMI-related hospitalizations were reduced by almost 50% during the COVID-19 period and accompanied by a 3-fold increase in mortality and complications (De Rosa et al 2020).

To the best of our knowledge, such papers are mostly observational studies, showing descriptive analysis (correlations, relative risks or odds ratio) or focusing on mortality trends for no-COVID patients (Birkmeyer et al. 2020), reporting de facto results before and after the Covid period, after having established some significant cutoff date (first case, lockdown etc).

However, there is still a lack of evidence assessing if Covid-19 and its related stress at hospital level has modified admission criteria and related mortality rates in a causal inference framework using quasi-experimental methods when there is potential endogeneity on comparison.

Moreover, other factors from the Covid-19 pandemic could affect patient behavior, such as seasonality of the assessed periods. In this end, some US (Salomon et al 2020) and Italian studies (De Filippo et al, 2020) demonstrates that the fall of incidence of hospitalizations for AMI during the first wave of Covid-19 pandemic, declined more than expected by typical seasonal variation alone.

The aim of the present paper is to assess if the extra-burden on hospital and staff (in one hospital in the epicenter of the pandemic in Italy) caused by Covid-19 pandemic has had consequences on intra-hospital mortality for patients admitted for no-Covid urgent conditions. Beside mortality, we also assess if the case-mix of hospitalized patients changed during the epidemic with respect to non-pandemic period both in a short and long-term perspective. The analysis focused on the first wave of the pandemic only allowing us to observe the very first reaction of the healthcare system to this unexpected event.

Methodologically we use regression discontinuity design (RDD), paired with difference-in-regression-discontinuity design (DRD) to get credible measures of the plausibly causal effects of the extra-burden on hospital and staff on intra-hospital mortality for patients admitted for no-Covid urgent conditions (AMI and stroke).

Data refer to hospitalizations of patients admitted at “Spedali Civili”, the main hospital in the province of Brescia, one of the most hit provinces in Italy by Covid-19. In fact, as reported by Italian Statistic Institute, in March 2020 this province experienced an overall mortality increase of 292% compared with the average of the same month in 2015-2019, and by the end of April it had registered 2,500 confirmed COVID-19 deaths (ISTAT 2020).

In particular, we select hospitalization records of patients admitted for acute stroke and acute myocardial infarction (AMI), comparing case mix and mortality rates across admissions during the first three months of the pandemic (since the lockdown date of March, 9^th^ until late spring 2020) both in an RDD design and also controlling for the same calendar period of no-Covid years in a DRD design.

The proposed analysis sheds light on potential effects of organizational and clinical practice changes to face Covid-19 in term of assuring high quality care for non-deferrable acute admissions in the Covid period.

To this end, it is important to note that Spedali Civili has implemented specific operational protocols to deliver an appropriate hospital care of no-Covid-19 emergency cases in the context of the first wave of the COVID-19 pandemic.

In fact, a progressive increase in Covid-19-devoted beds, either non-ICU or ICU-specific, rapidly reached around 800 beds out of a total of 1547 beds at the end of March 2020 (see Figure 1 in Appendix).

To face this situation, ER admissions were structurally modified in a **fully dual track system** by introducing a Covid-19-devoted triage and building external emergency tents to admit Covid-19 patients only. Spedali Civili was then literally transformed into a Covid-19 Hospital-hub meaning that a drastic modification has been realized both at structural and, most importantly, at organizational level: the already existing staff has been primarily involved in handling the emergency. Physicians, nurses and sanitary workers from both Infectious Diseases, ICU, and Pneumology wards were mainly involved, and received a specific training on Covid-19 management, also Internal Medicine doctors, Cardiologists, Neurologists, Surgeon, and Immunologists together with the related nurses, were also active part of Covid-19 patient care delivery.

However, despite the general prioritization of staff and resources on Covid-19 patients, for some time-dependent conditions (such as stroke, cardiovascular emergencies, neurosurgical emergencies, and trauma) an organization based on a “hub-and-spoke” model has been adopted and Spedali Civili was selected as the main regional “hub” also for Ami and Stroke cases in the eastern part of the Lombardy region. Requirements for being selected as “hub” included the “*presence of an integrated trauma team 24/7 on active duty and supplementary surgical teams available on call, fast-track access to Emergency Department to reduce interpersonal contact between patients, activation of separated pathways to assist and operate on COVID-19 and non-COVID-19 patients, and integration of local medical teams with those of the spoke centres*” (see Casiraghi et al. 2020 for more details).

## Methods

The paper is an observational retrospective, pre- and post-implementation study using administrative data from an important hospital set in the epicenter of the Lombardy region, where the Covid-19 epidemic has had a relevant impact in March-May 2020.

The main goal is to investigate the impact of Covid-19 pandemic (and related mitigation measures, such as lockdown) on patients’ mortality for AMI and stroke acute hospital admissions.

A classical causal inference method aims at estimating credible causal effects of treatments or policies in a quasi-random framework, when randomized controlled trials (RCTs) are not possible or not ethically/practically feasible.

Quasi-experimental techniques are very useful in this context and may provide a robust tool to draw information on causal impacts. To this end, the regression discontinuity design (RDD, Hahn et al. 2001) is one such quasi-experimental method that takes advantage of clinical or policy decision rules in which people are differentially assigned to a treatment or intervention if they fall above or below an arbitrary cut-off value of a continuous variable.

Causal inference within an RD framework comes from the assumption that, aside from differential use of treatment, those on either side - yet close to the cut-off are otherwise similar, and validity of the approach relies on the hypothesis that patients on either side of the threshold have comparable characteristics (as in a pure randomized study).

We take advantage of the strict cutoff date for the introduction of the Covid-19 lockdown in Italy, which was March 9^th^ 2020, that naturally divides the population of hospitalized patients into a treatment group composed by patients hospitalized after the cutoff date and a control group of patients hospitalized before the cutoff date.

In the RDD analysis patients in the treatment group, those admitted to the Spedali Civili of Brescia hospital with acute stroke or AMI during the first wave epidemic period (from March, 9^th^ to May, 31^st^ 2020) were compared to patients admitted before the lockdown implementation (January, 1^st^ to March, 8^th^ 2020) for the same diseases.

After having checked for observable differences in the covariates around the cutoff, similarly to Been et al (2020), we then use calendar week as running variable while the treatment status is identified adopting different time-windows around the cutoff. In particular, we assessed treatment effect by RDD in separate time windows (5 to 10 weeks before and after the cutoff date) assuming that any mortality change was due to the hospital-stress introduced by Covid-19 pandemic.

A second analysis was conducted to investigate differences in mortality due to the pandemic in an even more robust fashion. Although RDD design addresses the endogeneity of treatment in a quasi-experimental fashion, mortality differences by treatment (in the pandemic period) may be also induced by temporal trends or seasonal factors (i.e. winter vs spring) or by other underlying time-variant factors affecting mortality around the cutoff date. To this end, in the spirit of the difference in difference estimator, we compare mortality in the period surrounding implementation of the measures in 2020 to the same time periods in a year preceding the Covid-19 9 pandemic (pooling 2018-2019). More explicitly, mortality differences among treated (March, 9^th^ to May, 31^st^ 2020) and control patients (January, 1^st^ to March, 8^th^ 2020) were compared with differences in mortality of patients admitted during the corresponding periods in 2018-2019, when no restrictions were in place, and no stress due to the Covid-19 pandemic affected the hospitals activity.

In this end, we adopt a difference-in-regression-discontinuity design (DRD, Hong et al 2017 and Been et al. 2020) which identifies the effect of Covid-19 pandemic on mortality as the difference between the estimated effects on mortality of an RDD around the lockdown date (March,9^th^) in the year of Covid-19 expansion (2020) with the ones of an RDD around the same date in the pooled period 2018-2019, to control for pre-existing (observed or unobserved) differences in mortality determinants around the cutoff.

The causal impact, assessed within several time-windows (from 4 to 10 weeks) around the lockdown date (March 9^th^), was estimated with a logistic regression that models the probability of death at patient level as a function of a treatment dummy (assuming value of 1 after the cutoff date and 0 before the cutoff date), a cohort dummy (1 for cohort 2020 and 0 for cohorts 2018-2019), its interaction and other case-mix covariates to control for unbalanced case-mix among cohorts or among treated and controls in 2020.

The causal effect of DRD is identified as the interaction parameter, whose exponential value can be interpreted as the odds of mortality difference post/pre lockdown various time windows following implementation of the COVID-19 lockdown (March, 9th 2020) versus the odds of the same event across similar time windows (around the lockdown) in previous years (2018–2019) without the pandemic.

Data are gathered from the administrative information system of Spedali Civili of Brescia and collect information on patients admitted from January 2018 to May 2020. The hospital discharge data contains basic demographic information (age, gender), information on hospitalization (length of stay, special-care unit use, transfers within the same hospital or through other facilities, and in-hospital mortality), and a total of 6 diagnosis codes and procedures defined according to the International Classification of Diseases, Ninth Revision, Clinical Modification (ICD-9-CM).

The analysis was limited to the following ICD-9-CM codes detected as the principal diagnosis: 410 (Acute myocardial infarction) and 434 (Occlusion of cerebral arteries - Stroke). We exclude all discharges including as principal diagnosis the code 410.9, corresponding to an AMI with unspecified site. Patients with a principal diagnosis coded 410.7 are detected as AMI with Non-ST elevation myocardial infarction (NSTEMI), while the other 410 codes indicate an AMI with ST elevation myocardial infarction (STEMI).

A set of selected variables at patient level were chosen to control for determinants of patient mortality that may be used in the RDD and DRD model in case of unbalanced case-mix of treated and control group. In particular, at the patient level we control for patient’s age (in years); gender, foreign status and coexisting conditions identified by the Elixhauser algorithm (Elixahuser, 1998).

## Results

Data refers to 525 hospitalizations in 2020 (81% between March 9^th^ – May 31^st^ 2020, 18% in the pre-lockdown period January, 1^st^ – March, 9^th^), and 996 in 2018-2019

Table 1 reports the trend in the number of patients admitted at the hospital with a main diagnosis of AMI (both STEMI and NSTEMI), as well as the trend of patients’ mortality for the same time window (January-May) over the last three years.

**Table 1:** Mortality rates of AMI by type (STEMI and NSTEMI) (March 9-May 31)

| Year | STEMI |  |  | NSTEMI |  |  | TOT |  |  |
| --- | --- | --- | --- | --- | --- | --- | --- | --- | --- |
|  | Mean | Std.Dev | N | Mean | Std.Dev | N | Mean | Std.Dev | N |
| 2018 | 0.0750 | 0.2650 | 80 | 0.0363 | 0.1880 | 110 | 0.0526 | 0.2238 | 190 |
| 2019 | 0.0978 | 0.2987 | 92 | 0.0166 | 0.1285 | 120 | 0.0518 | 0.2223 | 212 |
| 2020 | 0.0743 | 0.2634 | 121 | 0.0312 | 0.1753 | 64 | 0.0594 | 0.2371 | 185 |

Overall, the number of AMI cases does not change dramatically over time, but we observe that the composition in terms of STEMI and NSTEMI changes in the last year if compared to 2018-2019. In 2018 and 2019 STEMI represents the 40% of the AMI admitted at Spedali Civili, while in 2020 the number of STEMI rises to 65% of the total. Suggestive evidence points out how this difference in AMI composition may be driven by the local healthcare system, which funneled many STEMI cases from decentralized hospitals to the Spedali Civili, which is the main hospital in the Brescia province.

In addition, we observe an almost steady (5%) total mortality rate for patients affected by AMI over the three years considered. These first descriptive comparison points towards a lack of significant change in volumes of total admissions and mortality (overall), suggesting an adequate response for this kind of time-depending and high-risk clinical condition, despite the pandemic exposed the hospital to an overwhelmed additional and unforeseen stress.

Differently, Table 2 exhibits a moderate increase in both volumes and mortality rates for Stroke cases among 2019 and 2020 with a large growth for both outcomes in 2018 and 2020.

**Table 2:**
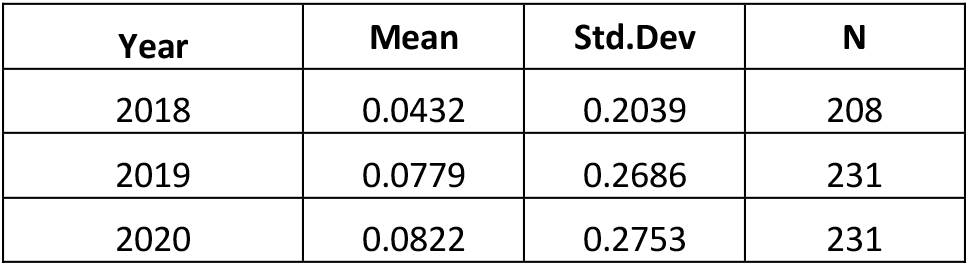
Mortality rates of Stroke (March 9-May 31)

In the first analysis we focused on the comparison of mortality around the lockdown date (March, 9^th^ 2020) in 2020. Figure 1 reports the observed weekly mortality rates (15-days windows) around the cutoff. It evidences a steady path in mortality after the threshold for AMI patients and a reversed U-shaped pattern for stroke patients.

**Figure 1:**
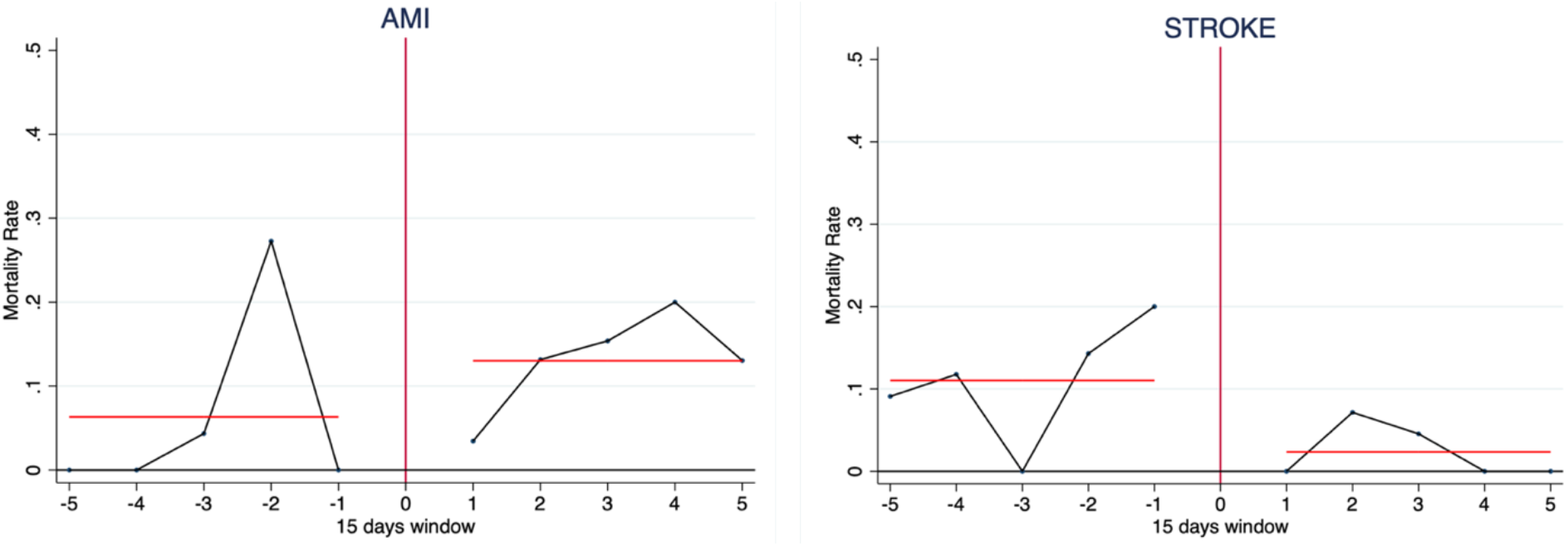
Observed 15-days windows mortality rates around the lockdown cutoff (March, 9th 2020). The red lines identify the average mortality in the pre and post lockdown periods

Fig 2 reports the estimated causal effect of the Covid-19 pandemic around the implementation date of the State of Emergency restrictions in the RDD model. Results show a non-statistically significant effect of the lockdown restrictions and related Covid-19 extra burden on hospitals activities on both AMI and Stroke mortality rates in all the 6 different time windows considered around the lockdown date (from 5-weeks to 10 weeks). The RDD logistic equation includes also covariates that are not balanced around the cutoff for the largest study period of 10 weeks.

**Figure 2:**
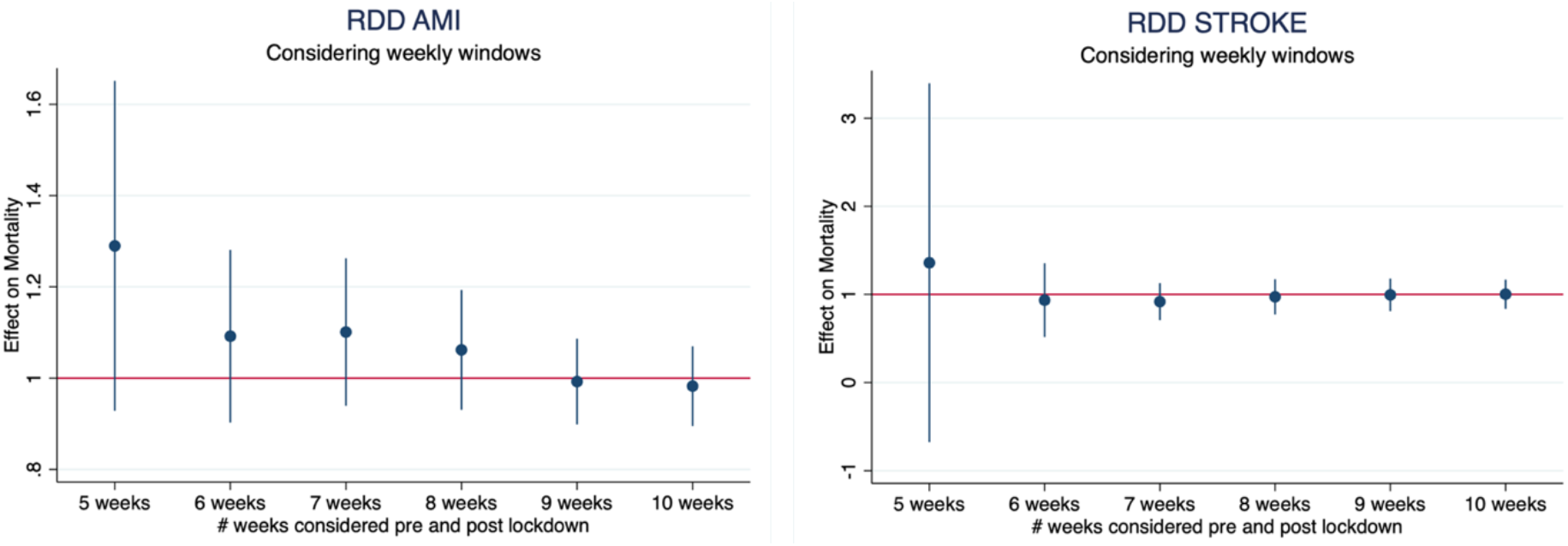
Odds ratios (95% CI) indicating odds of mortality across various time windows following implementation of the COVID-19 lockdown (March, 9^th^ 2020) versus the odds of mortality in similar time windows preceding the measures in 2020. Estimates derived from regression-discontinuity design.

The two groups are well balanced across covariates except for few case-mix variables and Comorbidities (calculated adopting the Elixhauser algorithm), such as length of hospital stay (increased after the cutoff) and presence of Peripheral Vascular Disorders (whose prevalence decrease after the cutoff) for AMI and Diabetes Complicated (whose prevalence decreases, although in a not significant manner) for stroke patients (See Table 3 and 4 in the Appendix).

As well as in the RDD case, Figure 3 checks the plausible causal effect of the Covid-19 pandemic over several time-windows of 15-days around the implementation date of the State of Emergency restrictions for COVID-19 on March, 9^th^. In particular, extending the bandwidth around the selected cutoff point, we check whether the impact of the additional stress for the healthcare system due to the Covid-19 pandemic has had an effect on the mortality rate of AMI and Stroke hospitalized patients. Results of the estimate causal effects provided by the regression-in-discontinuity model with their confidence intervals (at 95%) are then reported in Figure 3.

**Figure 3:**
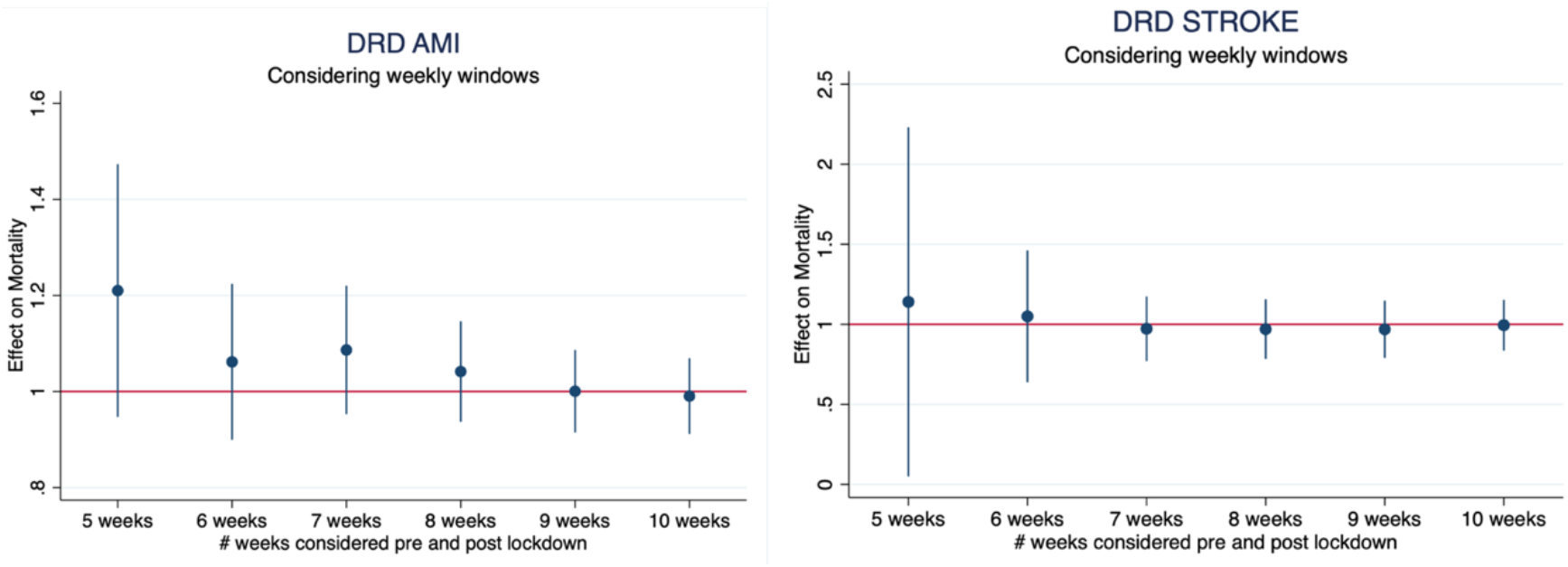
odds ratio (95% CI) indicating odds of mortality difference post/pre lockdown in various time windows following implementation of the COVID-19 lockdown (March, 9^th^ 2020) versus the odds of the same event in the similar time windows in previous years (2018–2019). Estimates derived from difference in regression-discontinuity design. DRD

Comparison among 2020 and 2018-2019 cohorts’ characteristics revealed a similar composition of hospitalizations for both AMI and Stroke with some exception: for AMI patients are slightly younger in 2020 (67.7 vs 70.1), and the quota of NSTEMI significantly decreases in 2020 (33.8% vs 56.2%). Prevalence of comorbidities varies significantly among cohorts for Cardiac Arrhythmias (increases at 22.5% from 10.1%), Peripheral Vascular Disorders (increases at 5.6% from 1.6%), and Other Neurological Disorders (increased at 1.9% from 0.2%).

As for stroke cases, cohorts are largely balanced for demographics and comorbidities, except for prevalence of patients affected by Congestive Heart Failure (that falls at 0.46% in 2020 from 2.8% in 2018-19). See Tables 5 and 6 in the Appendix for details.

Results of DRD (Figure 3), which controls for unobserved and seasonal effects, confirms RDD results: odd ratios are still non-significant, but also slightly closer to one in their magnitude at the different time-windows considered, when compared with RDD odd ratios.

For both analyses, particularly for AMI patients, the largest (although not significant) increase of mortality post the cutoff was found in the shortest time windows of 5 weeks around the lockdown (OR=1.21 for DRD), where the analyzed hospital has probably experienced the maximum organizational stress due to Covid-19 admissions. Moreover, such differences in mortality decreased regularly over time (windows) demonstrating that mortality rates in the Covid-19 period returned to pre-Covid-19 levels 9 weeks later than the lockdown, thus presumably at mid-May. Reaction to Covid stress for stroke seemed faster. Mortality rates in the Covid period returned to pre-Covid levels, six weeks later than lockdown (end of April).

Hence, results agree that - even if the extra-burden faced during the lockdown period has changed many of the ordinary activities of the hospital-acute AMI and Stroke mortality has not been affected by Covid-19 stress of the healthcare system in the long run. This is an important result demonstrating how the mitigation strategies put in place by the Spedali Civili of Brescia simultaneously face Covid-19 and no-Covid-19 healthcare delivery has been successfully implemented.

Before discussing the obtained estimates, it is important to note that - as pointed out in some recent contributions in the literature-Lombardy region has experienced - on average-an increase in the time from the onset of the first symptoms to the patients hospitalization during the pandemic and we know from the literature how early recognition of symptoms along with shorten time to intervention usually results in better outcomes following both stroke and AMI (Levine at al. 2013; Lees et al. 2010).

Unfortunately, times from the onset of patients’ symptoms to door and to balloon are still not available for the whole sample of hospitalizations under study. However, we were able to retrieve information on a subsample composed by 164 patients hospitalized in the 2020 first lockdown period compared with patients hospitalized in the same period in 2019 years. Table 3 offers descriptive statistics of the mean time occurred from the onset of first symptoms to Door and to Balloon as well as of the within hospital time between Door and Balloon. We then registered a statistically significant (at 10% level of confidence only) increase of 2 hours and 51 mins on average from the onset of symptoms to door and a non-significantly different from 2019 but larger time from door to balloon of 18 mins - which may be du to Covid-19 testing procedures at the hospital admission stage-in 2020.

**Table 3:** Descriptive statistics of AMI time from Onset Symptoms by year (2019 vs 2020)

| Year | N. Obs | Onset Symptoms to Door |  | Door to Balloon |  | Onset Symptoms to Balloon |  |
| --- | --- | --- | --- | --- | --- | --- | --- |
|  |  | Mean | Std Error | Mean | Std Error | Mean | Std Error |
| 2019 | 64 | 206 | 30.13 | 77.68 | 11.3 | 254.72 | 28.45 |
| 2020 | 100 | 377.12 | 85.41 | 95.82 | 11.11 | 476.31 | 88.7 |
| 2019-2020 |  | -171.12 | 90.57 | -18.13 | 15.86 | -221.587 |  |
| t-stat |  | -1.88 |  | -1.14 |  | -2.37 |  |
| p-value |  | 0.0615 |  | 0.2547 |  | 0.0192 |  |

On average a total increase of the global time from the onset of symptoms to Balloon is around 3 hours and 41 mins (statistically significant at a 5% confidence level) in line with recent study at Humanitas Hospital in Lombardy (see Asteggiano et al., 2021).

A similar pattern has been experienced also for stroke episodes with an average increase of 10 mins-from 126 mins in 2019 to 136 mins in the first half of 2020-in the onset of symptoms to door time computed using all direct in-hospital admissions via ambulance.

Given the registered increase of these times we would a priori expect a potentially negative effect on mortality.

Unfortunately, it is not possible to formally consider the time-to-hospitalization as a covariate in our models given the availability of this information only at aggregated level for stroke and for a subsample of 164 patients only for AMI. However, despite of the potential adverse effect on mortality expected due to a longer door-to-needle time the AMI and Stroke mortality rates in our quasi-experimental setting are not statistically different from the one observed in the control groups. These non-significant difference in mortality between the great 2020 lockdown period and the same period in 2019 may be the results of a better management of AMI and Stroke cases given the implemented dual-track organizations. Having a dedicated A&E admission department and a 24/7 dedicated team of doctor and nurses as well as an ICU available only for these type of emergency admissions may have contributed to lowering the negative effects of longer times to hospitalization.

## Conclusions

To our knowledge, our study is the first to have assessed the impact of COVID-19 and related burden on hospital stress on in-hospital mortality for AMI and STROKE in one of the largest Northern Italian Hospital sited in the Province of Brescia (the main pandemic center in Europe in the first wave, recorded 2,500 confirmed COVID-19 deaths by the end of April) in a causal inference framework.

In this end, quasi-experimental evidence resulted useful to complement the descriptive evidence brought about by the standard observational studies on AMI and STROKE care delivery in Covid-19 times.

A regression discontinuity design was implemented comparing the weekly mortality rates difference after and before the cutoff point (lockdown date) and complemented with a difference in regression discontinuity fashion (difference occurred across the same cutoff in 2018-2019) to assess the Covid-19 effect, controlling for seasonal and other unobserved factors that were possibly in place when no pandemic was in place.

Both RDD and DRD demonstrates that, although we expected illness severity in non-Covid-19 admissions to increase (due to possible selection criteria that could have avoided fewer sick patients) in-hospital mortality in patients without COVID-19 was not different during the first wave of the pandemic with respect to pre-lockdown period.

Mortality rates in the Covid-19 period returned to pre-Covid-19 levels, nine and six weeks later than lockdown for AMI and Stroke patients, respectively.

Concerning admission criteria of hospitalization in two analyzed periods (2020 vs 2018-2019), patients’ characteristics of hospitalized are largely similar, with some exceptions (Cardiac Arrhythmias and Peripheral Vascular Disorders, which are significantly increased in 2020 for AMI patients, unlikely a large reduction in NSTEMI patient). A similar decreasing pattern was observed for Congestive Heart Failure patients admitted for stroke.

Results show that no-Covid cases (in particular, non-deferrable clinical issues) can be effectively managed adopting a fully dual track system in the hospital setting which helps in avoiding that the unexpected extra-stress due to Covid-19 hospitalizations would compromise the quality of cares to provided.

Our study also has limitations. First, the proposed evidence is limited to one single hospital and this might not reflect the clinical reality at every hospital in Northern Italy during the first wave of the pandemic. Second, we measured in-hospital mortality only and therefore we are unable to quantify either mortality for patients affected by AMI or stroke but not hospitalized and mortality post-discharge (i.e. 30 days mortality). Third, our analysis reflects less than a full year of data and ends in later May 2020. Availability of similar data for the regional context would be helpful in further substantiate the obtained findings and in exploring existing differences in efficacy of different managerial settings implemented in Lombardy hospitals.

## Data Availability

The authors are open to sharing statistical codes. Agreement of the Spedali Civili of Brescia, the data provider, will be required for any data sharing.

## Acknowledgments

Opinions and views expressed in this paper are those of the authors and do not necessarily reflect the ones of the institutions affiliation.

## Data availability

The authors are open to sharing statistical codes. Agreement of the “Spedali Civili of Brescia”, the data provider, will be required for any data sharing.

## Author Contributions

### Conceptualization

P. Berta, S. Curello, P. G. Lovaglio, M. Magoni, M. Metra, A. M. Roccaro, C. Rossi, S. Verzillo, G. Vittadini

### Data Analysis

P. Berta and P. G. Lovaglio

### Writing-review & editing

P. Berta, S. Curello, P. G. Lovaglio, M. Magoni, M. Metra, A. M. Roccaro, C. Rossi, S. Verzillo, G. Vittadini

## Author approval

All authors have seen and approved the manuscript.

## Competing Interests

No competing interests are declared by the authors

## Appendix

**Figure 1:**
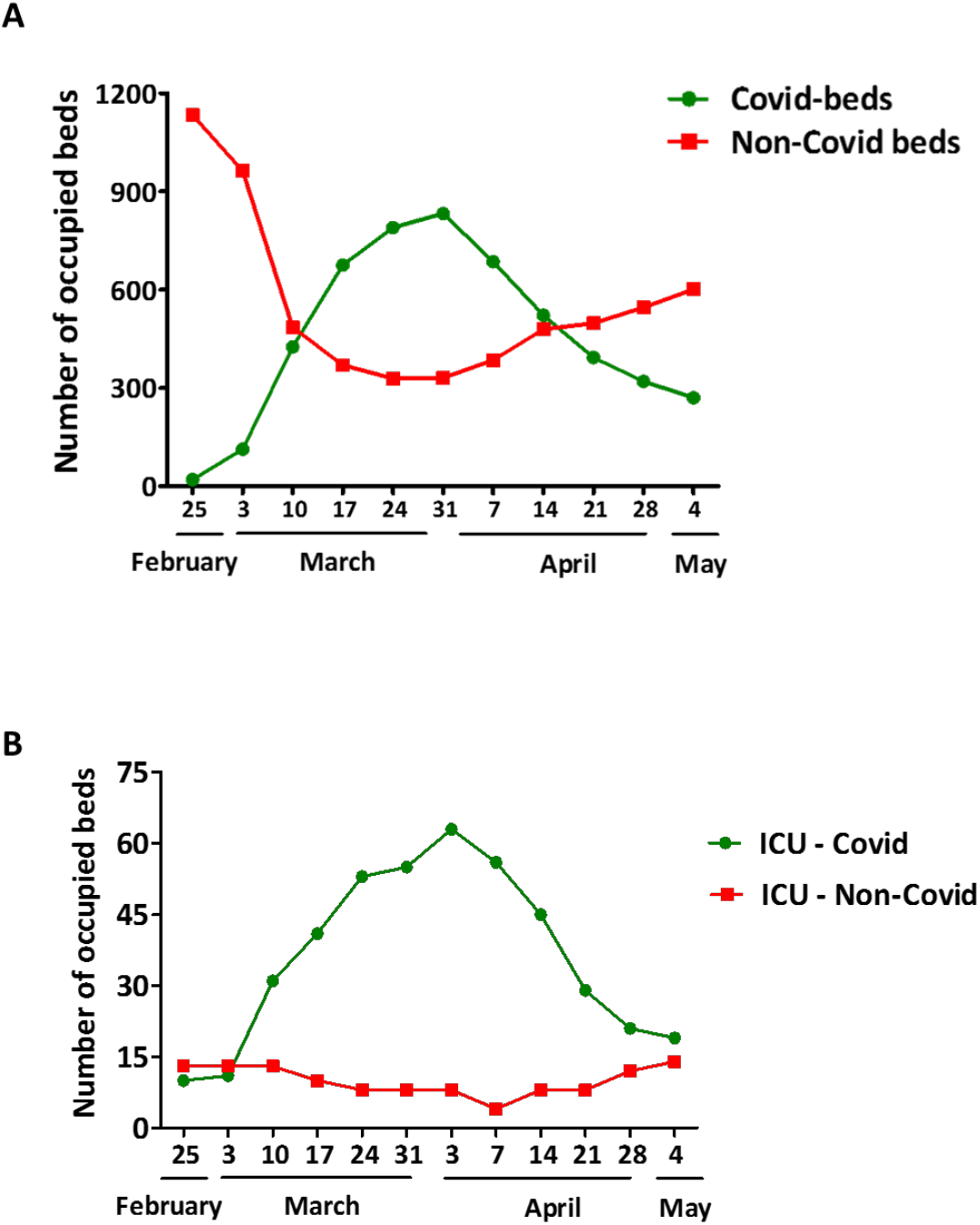
Descriptive statistics of Covid-29 and non-Covid-19 occupied beds in ordinary (Panel A) and acute/ICU wards (panel B) during the first pandemic wave

**Table 3:** Differences of AMI patients characteristics pre and post lockdown (March, 9^th^) in 2020

| Variables | Pre | Post | p value |
| --- | --- | --- | --- |
| Age | 66.214 | 68.065 | 0.5874 |
| Female | 0.214 | 0.271 | 0.6482 |
| NSTEMI | 0.429 | 0.342 | 0.5177 |
| Length of stay | 4.929 | 7.658 | **0.0212** |
| Foreign | 0.214 | 0.065 | 0.0443 |
| Congestive Heart Failure | 0.000 | 0.097 | 0.2251 |
| Cardiac Arrhythmias | 0.071 | 0.226 | 0.1787 |
| Valvular Disease | 0.143 | 0.058 | 0.2205 |
| Pulmonary Circulation Disorders | 0.000 | 0.019 | 0.6020 |
| **Peripheral Vascular Disorders** | **0.357** | **0.058** | **0.0001** |
| Hypertension, Uncomplicated | 0.000 | 0.006 | 0.7648 |
| Other Neurological Disorders | 0.000 | 0.019 | 0.6020 |
| Diabetes, Uncomplicated | 0.000 | 0.026 | 0.5458 |
| Hypothyroidism | 0.000 | 0.006 | 0.7648 |
| Renal Failure | 0.000 | 0.032 | 0.4980 |
| Solid Tumor Without Metastasis | 0.000 | 0.006 | 0.7648 |
| Coagulopathy | 0.000 | 0.006 | 0.7648 |
| Obesity | 0.000 | 0.006 | 0.7648 |
| Fluid and Electrolyte Disorders | 0.000 | 0.006 | 0.7648 |
| Blood Loss Anemia | 0.000 | 0.013 | 0.6712 |
| Deficiency Anemia | 0.000 | 0.013 | 0.6712 |
| Hypertension, Complicated | 0.000 | 0.019 | 0.6020 |

**Table 4:**
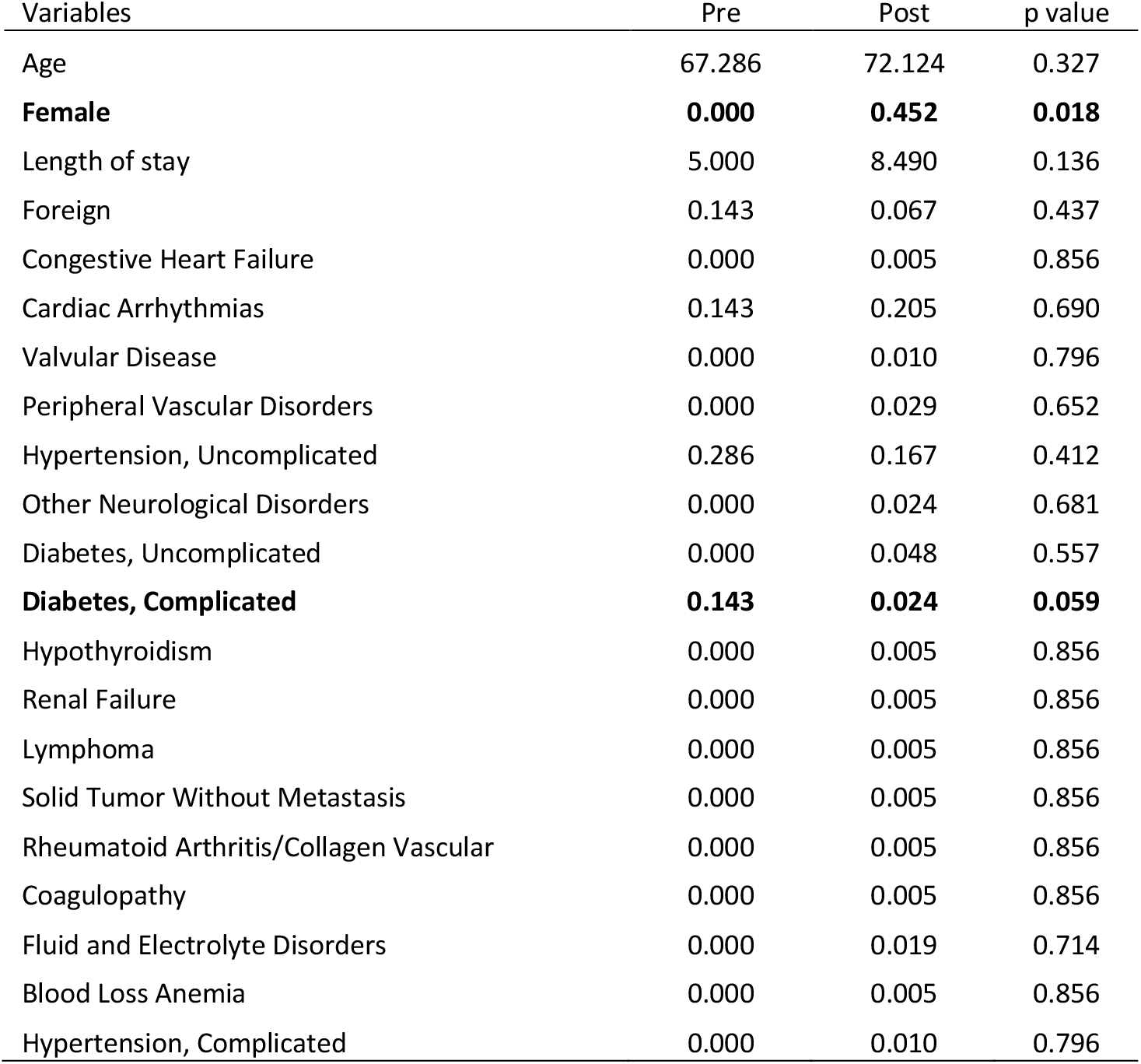
Differences of Stroke patients characteristics pre and post lockdown (March, 9^th^) in 2020

**Table 5:** Differences of AMI patients characteristics between 2020 and 2018/19

| Variable | 2018-19 | 2020 | p value |
| --- | --- | --- | --- |
| **Age** | **70.0890** | **67.7625** | **0.0489** |
| Female | 0.3044 | 0.2688 | 0.3991 |
| **Nstemi** | **0.5621** | **0.3375** | **0.0000** |
| Foreign patient | 0.0679 | 0.0625 | 0.8149 |
| Congestive Heart Failure | 0.1124 | 0.1000 | 0.6681 |
| **Cardiac Arrhythmias** | **0.1007** | **0.2250** | **0.0001** |
| Valvular Disease | 0.0375 | 0.0563 | 0.3165 |
| Pulmonary Circulation Disorders | 0.0070 | 0.0188 | 0.2092 |
| **Peripheral Vascular Disorders** | **0.0164** | **0.0563** | **0.0082** |
| Hypertension, Uncomplicated | 0.0141 | 0.0063 | 0.4390 |
| **Other Neurological Disorders** | **0.0023** | **0.0188** | **0.0314** |
| Chronic Pulmonary Disease | 0.0094 | 0.0000 | 0.2200 |
| Diabetes, Uncomplicated | 0.0141 | 0.0250 | 0.3622 |
| Diabetes, Complicated | 0.0023 | 0.0000 | 0.5409 |
| Hypothyroidism | 0.0000 | 0.0063 | 0.1024 |
| Renal Failure | 0.0281 | 0.0313 | 0.8399 |
| Liver Disease | 0.0023 | 0.0000 | 0.5409 |
| Lymphoma | 0.0023 | 0.0000 | 0.5409 |
| Solid Tumor Without Metastasis | 0.0047 | 0.0063 | 0.8131 |
| Coagulopathy | 0.0023 | 0.0063 | 0.4702 |
| Obesity | 0.0023 | 0.0063 | 0.4702 |
| Fluid and Electrolyte Disorders | 0.0023 | 0.0063 | 0.4702 |
| Blood Loss Anemia | 0.0117 | 0.0125 | 0.9375 |
| Deficiency Anemia | 0.0117 | 0.0125 | 0.9375 |
| Hypertension, Complicated | 0.0047 | 0.0188 | 0.0990 |

**Table 6:** Differences of Stroke patients characteristics between 2020 and 2018/19

| Variable | 2018-19 | 2020 | p value |
| --- | --- | --- | --- |
| Age | 70.3786 | 72.2019 | 0.1185 |
| Female | 0.4486 | 0.4507 | 0.9590 |
| Foreign patient | 0.0547 | 0.0657 | 0.5711 |
| Congestive Heart Failure | 0.0284 | 0.0047 | 0.0454 |
| Cardiac Arrhythmias | 0.1729 | 0.2113 | 0.2339 |
| Valvular Disease | 0.0328 | 0.0094 | 0.0727 |
| Pulmonary Circulation Disorders | 0.0066 | 0.0000 | 0.2366 |
| Peripheral Vascular Disorders | 0.0438 | 0.0282 | 0.3312 |
| Hypertension, Uncomplicated | 0.1926 | 0.1690 | 0.4656 |
| Paralysis | 0.0044 | 0.0000 | 0.3343 |
| Other Neurological Disorders | 0.0241 | 0.0235 | 0.9626 |
| Chronic Pulmonary Disease | 0.0066 | 0.0000 | 0.2366 |
| Diabetes, Uncomplicated | 0.0656 | 0.0469 | 0.3423 |
| Diabetes, Complicated | 0.0197 | 0.0282 | 0.4906 |
| Hypothyroidism | 0.0044 | 0.0047 | 0.9542 |
| Renal Failure | 0.0175 | 0.0047 | 0.1803 |
| Liver Disease | 0.0022 | 0.0000 | 0.4952 |
| Lymphoma | 0.0022 | 0.0047 | 0.5804 |
| Solid Tumor Without Metastasis | 0.0109 | 0.0047 | 0.4250 |
| Rheumatoid Arthritis | 0.0066 | 0.0047 | 0.7703 |
| Coagulopathy | 0.0066 | 0.0047 | 0.7703 |
| Obesity | 0.0022 | 0.0000 | 0.4952 |
| Fluid and Electrolyte Disorders | 0.0328 | 0.0188 | 0.3086 |
| Blood Loss Anemia | 0.0022 | 0.0047 | 0.5804 |
| Deficiency Anemia | 0.0066 | 0.0000 | 0.2366 |
| Psychoses | 0.0022 | 0.0000 | 0.4952 |
| Depression | 0.0022 | 0.0000 | 0.4952 |
| Hypertension, Complicated | 0.0306 | 0.0094 | 0.0938 |

## Notes

### Competing Interest Statement

The authors have declared no competing interest.

### Author Declarations

This is to confirm that the study titled "The impact of COVID-19 on AMI and Stroke mortality in Lombardy: Evidence from the epicenter of the pandemic", authored by "Rossi C., Berta P., Curello S., Lovalgio P.G., Magoni M., Metra M., Roccaro A.M., Verzillo S., Vittadini G.", does not require an ethical approval. Sincerely, Sandra Sigala Brescia Ethics Committee President

